# Heat wave, COVID-19, and mortality excess in the 2022 summer: a cohort study on data from Italian surveillances

**DOI:** 10.1101/2022.12.12.22283336

**Authors:** Francesco Venturelli, Pamela Mancuso, Massimo Vicentini, Marta Ottone, Cinzia Storchi, Francesca Roncaglia, Eufemia Bisaccia, Chiara Ferrarini, Patrizio Pezzotti, Paolo Giorgi Rossi, The Reggio Emilia COVID19 Working group

**Affiliations:** Epidemiology Unit, Azienda USL – IRCCS di Reggio Emilia, Reggio Emilia, Italy; Public Health Unit, Azienda USL – IRCCS di Reggio Emilia, Reggio Emilia, Italy; Department of Infectious Diseases, Istituto Superiore di Sanità, Rome, Italy

**Keywords:** Mortality, Heatwave, COVID-19, Surveillance, Sars-CoV-2

## Abstract

We aimed to assess differences in the summer excess of mortality by COVID-19 history using data from the mortality and COVID-19 surveillances. We found 4% excess risk in 2022 summer, compared to 2015-2019. A mortality rate ratio of 1.59 (95%CI 1.39-1.82) for COVID-19 survivors compared to naïve, was found. Both were higher in people aged ≥75 years. During the July heat wave, the excess for COVID-19 survivors decreased and disappeared when excluding people living in nursing homes.

**Funding statement:** This study was partially supported by the Italian Ministry of Health –CCM 2020 - “Sorveglianza epidemiologica e controllo del COVID-19 in aree urbane metropolitane e per il contenimento della circolazione del Sars-CoV-2 nella popolazione immigrata in Italia” and by the Ricerca Corrente 2023

**Highlights:**

- the excess of mortality in COVID-19 survivors is not exacerbated by heatwaves
- an excess of mortality during the whole summer in COVID-19 survivors aged over 75 suggest that no harvesting effect is appreciable in the older population that survived COVID-19
- For COVID-19 survivors aged over 75, a lower mortality than the naïve population was observed only during the July heat wave when we stratified by residency

## Introduction

In 2022 summer, an excess of mortality was observed in many European areas, particularly among older people [1,2]. The excess was probably linked to the extremely high temperature observed during the 2022 summer. [1-3] The impact of heat waves on mortality is well known, affecting in particular, but not only, people with chronic conditions.[4-7] Furthermore, for the first time in 2022, an extreme heat wave occurred in a population with a large proportion of people who recovered from COVID-19.

COVID-19 had a large impact on mortality in 2020 [8], 2021 [9] and at the beginning of 2022 [10] in Italy, affecting in particular people with chronic conditions and possibly modifying the proportion of frail subjects in the population.[11] Therefore, the COVID-19 pandemic may have interacted with the heatwave in different ways. On one hand, the strong excess mortality in people with chronic disease may have reduced those susceptible to dying during the heat waves. On the other hand, having had COVID-19 could be itself a predisposing factor increasing susceptibility to heatwaves.[12]

We conducted an analysis to assess if the observed excess mortality in summer 2022 was stronger in COVID-19 survivors.

## Methods

### Study design and setting

Population-based cohort study based on surveillance data from the Reggio Emilia province, Italy.

### Statistical analysis and data sources

To assess the excess of mortality, we computed standardized mortality ratios (SMR) for the 2022 summer compared to mortality that occurred in previous summers (2015-2019, 2020, and 2021). SMRs were computed for the entire period (from June 15 to September 15) and for the periods with the major heatwaves in 2022 (July and the second fortnight of July). SMRs were computed for the whole population, adjusted by sex and age (5-year categories from 0 to 100 years) and stratified by sex and age (<75 and ≥75 years). These data come from the summer mortality surveillance [13] that collects aggregated data on daily mortality of the resident population occurring in all the municipalities of the province of Reggio Emilia, i.e. deaths occurring outside the province of Reggio Emilia were not included.

To assess differences in mortality by COVID-19 history, we computed the mortality rate ratios (MRR), adjusted for age and sex, of dying for any cause during the 2022 summer period. People who recovered from COVID-19 (namely COVID-19 survivors) were compared to never-infected individuals (namely naïve). COVID-19 history was classified according to data from the COVID-19 surveillance system and taking into account all the SARS-CoV-2 infections diagnosed between February 20, 2020 and May 31, 2022. All residents in the Reggio Emilia province on December 31, 2019, and alive on June 15, 2022, were included. People who had COVID-19 between June 1-14 2022 were excluded while people diagnosed between June 15 and September 15 were censored at the date of infection. We computed MRRs for the whole summer period and the same sub-periods considered for SMRs, for the entire population and stratified by sex and age. IRRs in people older than 74 years were also stratified by residency (nursing home vs community-dwelling residents) deduced by being tested or not for SARS-CoV-2 within nursing home routine screenings.

Finally, we used the time series analysis to explore the association between the Thom Discomfort Index [14] and the daily number of deaths, using a lag of 0, 1, and 3 days. The Thom Discomfort Index was built using daily average temperature and humidity. Time series analyses were stratified by COVID-19 history (COVID-19 survivors or naïve).

## Results and discussion

The resident population in the Reggio Emilia province on June 15 2022 was 522,668, of which 151,107 were COVID-19 survivors. A total of 1251 deaths occurred during the 2022 summer, of which 303 in COVID-19 survivors and 948 among naïve people. 14 and 150 deaths, respectively, occurred after a SARS-CoV-2 infection diagnosed between July 15, and September 15, 2022, and were therefore excluded from MRR analyses. The summer mortality surveillance reported a 4% excess of mortality in 2022, while in the summers of 2020 and 2021 there was no excess. The 2022 excess was particularly marked in the second fortnight of July. The excess was appreciable in both sexes, but not in people below 75 (Table 1).

**Table 1.** Standardized mortality ratios (SMR) for summer mortality in Reggio Emilia province, Italy, years 2020, 2021, 2022 compared to 2015-2019. 95% CI = 95% Confidence Interval.

|  | Mortality in the summer period<br>(from June 15 to September 15) |  |  |  |  | Mortality in July<br>(from July 1 to July 31) |  |  |  |  | Mortality in the last fortnight of July<br>(from July 16 to July 31) |  |  |  |  |
| --- | --- | --- | --- | --- | --- | --- | --- | --- | --- | --- | --- | --- | --- | --- | --- |
|  | Observed | Expected | SMR | 95% CI |  | Observed | Expected | SMR | 95% CI |  | Observed | Expected | SMR | 95% CI |  |
| Overall |  |  |  |  |  |  |  |  |  |  |  |  |  |  |  |
| 2020 | 1240 | 1253 | 0,990 | 0,935 | 1,046 | 402 | 419 | 0,959 | 0,868 | 1,058 | 178 | 233 | 0,764 | 0,656 | 0,885 |
| 2021 | 1261 | 1255 | 1,005 | 0,950 | 1,062 | 437 | 418 | 1,045 | 0,949 | 1,147 | 216 | 232 | 0,929 | 0,810 | 1,062 |
| 2022 | 1315 | 1267 | 1,038 | 0,983 | 1,096 | 475 | 422 | 1,126 | 1,027 | 1,232 | 281 | 235 | 1,197 | 1,061 | 1,346 |
| Age stratified |  |  |  |  |  |  |  |  |  |  |  |  |  |  |  |
| 2020 |  |  |  |  |  |  |  |  |  |  |  |  |  |  |  |
| ≤74 | 254 | 266 | 0,954 | 0,841 | 1,079 | 85 | 91 | 0,935 | 0,747 | 1,156 | 40 | 55 | 0,731 | 0,522 | 0,995 |
| ≥75 | 986 | 987 | 0,999 | 0,938 | 1,063 | 317 | 328 | 0,966 | 0,863 | 1,079 | 138 | 178 | 0,774 | 0,650 | 0,914 |
| 2021 |  |  |  |  |  |  |  |  |  |  |  |  |  |  |  |
| ≤74 | 261 | 273 | 0,955 | 0,843 | 1,079 | 92 | 93 | 0,988 | 0,797 | 1,212 | 47 | 56 | 0,838 | 0,616 | 1,115 |
| ≥75 | 1000 | 982 | 1,019 | 0,956 | 1,084 | 345 | 325 | 1,061 | 0,952 | 1,179 | 169 | 176 | 0,958 | 0,819 | 1,114 |
| 2022 |  |  |  |  |  |  |  |  |  |  |  |  |  |  |  |
| ≤74 | 264 | 274 | 0,965 | 0,852 | 1,089 | 77 | 94 | 0,823 | 0,650 | 1,029 | 43 | 56 | 0,762 | 0,552 | 1,027 |
| ≥75 | 1051 | 993 | 1,058 | 0,995 | 1,124 | 398 | 328 | 1,212 | 1,096 | 1,337 | 238 | 178 | 1,334 | 1,170 | 1,515 |
| Sex stratified |  |  |  |  |  |  |  |  |  |  |  |  |  |  |  |
| 2020 |  |  |  |  |  |  |  |  |  |  |  |  |  |  |  |
| M | 582 | 590 | 0,986 | 0,908 | 1,070 | 193 | 197 | 0,979 | 0,846 | 1,127 | 91 | 112 | 0,816 | 0,657 | 1,001 |
| F | 658 | 663 | 0,992 | 0,918 | 1,071 | 209 | 222 | 0,942 | 0,819 | 1,079 | 87 | 121 | 0,716 | 0,574 | 0,883 |
| 2021 |  |  |  |  |  |  |  |  |  |  |  |  |  |  |  |
| M | 582 | 594 | 0,980 | 0,902 | 1,062 | 206 | 198 | 1,042 | 0,905 | 1,195 | 108 | 111 | 0,970 | 0,796 | 1,171 |
| F | 679 | 661 | 1,028 | 0,952 | 1,108 | 231 | 221 | 1,047 | 0,916 | 1,191 | 108 | 121 | 0,892 | 0,732 | 1,077 |
| 2022 |  |  |  |  |  |  |  |  |  |  |  |  |  |  |  |
| M | 617 | 605 | 1,020 | 0,941 | 1,104 | 233 | 201 | 1,160 | 1,015 | 1,318 | 136 | 114 | 1,195 | 1,002 | 1,413 |
| F | 698 | 662 | 1,054 | 0,978 | 1,136 | 242 | 221 | 1,096 | 0,962 | 1,243 | 145 | 121 | 1,199 | 1,012 | 1,411 |

MRRs showed an excess risk of death in COVID survivors compared to naïve people (MRR 1.59, 95%CI 1.39 to 1.82) (Table 2). The excess was stronger in females and people aged over 74, and it was appreciable in all the analyzed periods. In people aged over 74, the excess decreased when stratifying by residency (community-dwelling vs nursing home residents). Furthermore, the excess decreased when focusing the analysis on the period when the excess in the overall population was more marked and the temperature was extraordinarily higher, as in the second fortnight of July (MRR 1.20, 95%CI 0.88 to 1.65). In the same period, stratifying by residency the people aged over 74, the risk of dying was lower in COVID-19 survivors than in naïve people. (Table 2)

**Table 2.**
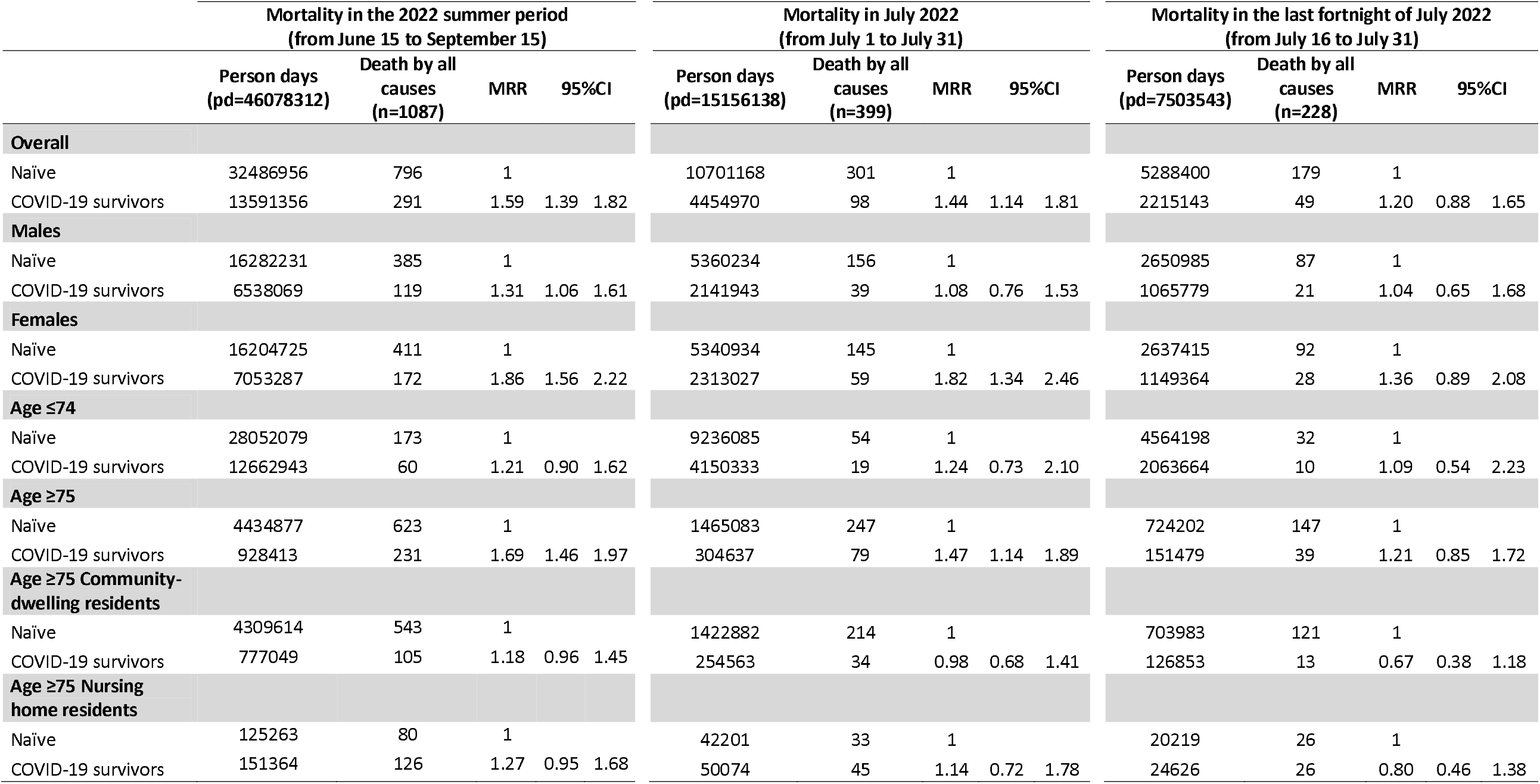
Poisson regression comparing the probability of dying for any cause in COVID-19 survivors and naïve people (assessed on May 31, 2022), during the 2022 summer in Reggio Emilia province Italy. Mortality Rate Ratios (MRR) are adjusted by age and sex. 95% CI = 95% Confidence Interval.

The time series analysis confirmed the association between temperatures and mortality in naïve people, showing a 8% excess (95%CI 2 to 13) for a one-degree increase of Thom Discomfort Index without any lag, rising to 10% and 8% with 1- and 3-days lags, respectively. While in COVID-19 survivors the effect was almost null with -1% (95%CI -9 to 9), 0% (95%CI -9 to 9) and 0% (95%CI -8 to 10) differences in mortality, with 0, 1- and 3-day lags respectively. (Table 3, Figure 1)

**Table 3.** Time series analysis for the association between Thom Discomfort Index (TDI) or maximum temperature (Tmax) and daily all-cause mortality in COVID-19 survivors and in COVID-19 naïve people (assessed on May 31, 2022). Summer 2022, Reggio Emilia, Italy.

|  | Covid-19 naïve |  | Covid-19 survivors |  |
| --- | --- | --- | --- | --- |
|  | Mortality rate ratio<br>per 1-degree<br>increase | 95%CI | Mortality rate ratio<br>per 1-degree<br>increase | 95%CI |
| TDI lag 0 | 1.08 | 1.02-1.13 | 0.99 | 0.91-1.09 |
| TDI lag 1 day | 1.10 | 1.04-1.15 | 1.00 | 0.91-1.09 |
| TDI lag 3 days | 1.09 | 1.03-1.14 | 1.00 | 0.92-1.10 |
| Tmax lag 0 | 1.02 | 1.00-1.04 | 1.02 | 0.98-1.06 |
| Tmax lag 1 day | 1.03 | 1.01-1.05 | 1.00 | 0.96-1.04 |
| Tmax lag 3 days | 1.04 | 1.01-1.06 | 1.00 | 0.96-1.04 |

**Figure 1.**
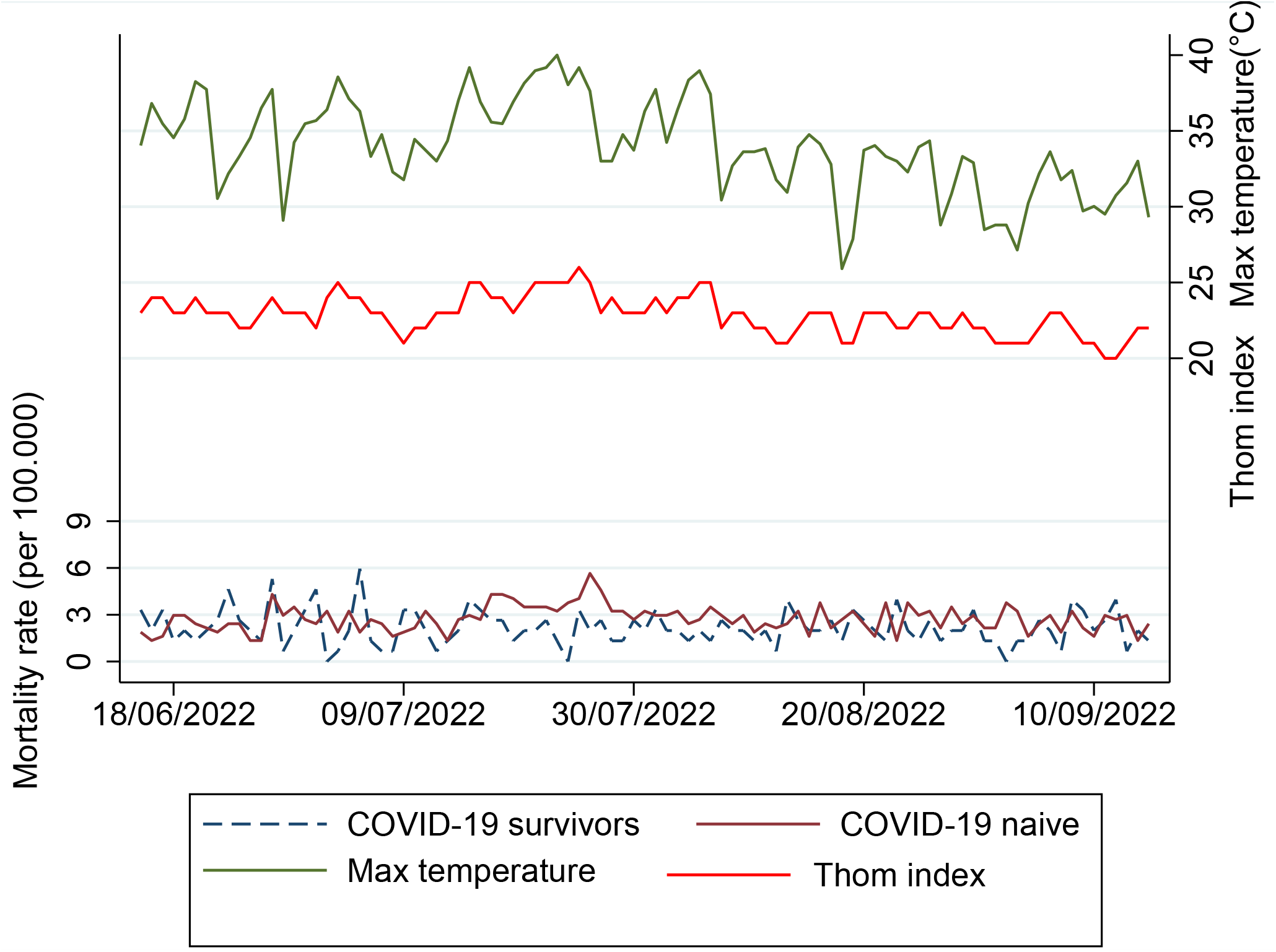
Daily mortality rate in COVID-19 survivors and COVID-19 naïve (assessed on May 31, 2022), Thom Discomfort Index and maximum Temperature in Reggio Emilia, Italy, 2022summer.

Our data showed that COVID-19 survivors have a strong excess of all-cause mortality as reported in a previous study from Estonia. [15]

We also confirm an excess mortality during the 2022 summer as already reported by several national statistics and we confirmed its link with the extremely high temperatures.[1-3] In our province, the 2022 summer excess of mortality was not appreciable in people below 75 years. A lower effect was expected, based on previous studies on the impact on mortality of heat waves, but not the absence of any excess.[16] This could be due to the small number of deaths in this age group, but also to a reduction of susceptible people due to previous COVID-19 mortality.

Our data suggest that the excess of mortality in COVID-19 survivors is not exacerbated by heatwaves. Furthermore, observing an excess of mortality during the whole summer in COVID-19 survivors aged over 75 also suggest that no harvesting effect is appreciable in the older population that survived COVID-19. The harvesting effect or mortality displacement is the occurrence of death earlier than it would have otherwise occurred; it implies that after an increase of deaths occurred for the harvesting phenomenon, a reduction in deaths would be observed. For COVID-19 survivors aged over 75, a lower mortality than the naïve population was observed only during the July heat wave when we stratified by residency. This result may suggest that a reduction of the susceptible population could have occurred and become appreciable only when exposed to an external stress factor.

## Data Availability

According to Italian law, anonymized data can only be made publicly available if there is no potential for the re-identification of individuals (https://www.garanteprivacy.it). Thus, the data underlying this study are available on request to researchers once collapsed on the patterns of covariates. Data access requests should be addressed to.

## References

1. ONS website, statistical article, Excess mortality during heat-periods: 1 June to 31 August 2022 Available at: https://www.ons.gov.uk/peoplepopulationandcommunity/birthsdeathsandmarriages/deaths/articles/excessmortalityduringheatperiods/englandandwales1juneto31august2022 Accessed on: November 28, 2022.

2. EuroMOMO Bulletin, Week 32, 2022. Available at: https://www.euromomo.eu/bulletins/2022-32 Accessed on: November 28, 2022.

3. WHO Europe. Statement – Climate change is already killing us, but strong action now can prevent more deaths. Statement by WHO Regional Director for Europe Dr Hans Henri P. Kluge. November 7, 2022. Available at: https://www.who.int/europe/news/item/07-11-2022-statement---climate-change-is-already-killing-us--but-strong-action-now-can-prevent-more-deaths. Accessed on: November 28, 2022.

4. Wu Y, Li S, Zhao Q, et al. Global, regional, and national burden of mortality associated with short-term temperature variability from 2000-19: a three-stage modelling study. Lancet Planet Health. 2022;6(5):e410–e421. doi:10.1016/S2542-5196(22)00073-0

5. Madaniyazi L, Armstrong B, Chung Y, et al. Seasonal variation in mortality and the role of temperature: a multi-country multi-city study. Int J Epidemiol. 2022;51(1):122–133. doi:10.1093/ije/dyab143

6. Zhao Q, Guo Y, Ye T, et al. Global, regional, and national burden of mortality associated with non-optimal ambient temperatures from 2000 to 2019: a three-stage modelling study. Lancet Planet Health. 2021;5(7):e415–e425. doi:10.1016/S2542-5196(21)00081-4

7. Bouchama A, Dehbi M, Mohamed G, Matthies F, Shoukri M, Menne B. Prognostic factors in heat wave related deaths: a meta-analysis. Arch Intern Med. 2007;167(20):2170–2176. doi:10.1001/archinte.167.20.ira70009

8. Dorrucci M, Minelli G, Boros S, et al. Excess Mortality in Italy During the COVID-19 Pandemic: Assessing the Differences Between the First and the Second Wave, Year 2020. Front Public Health. 2021;9:669209. Published 2021 Jul 16. doi:10.3389/fpubh.2021.669209

9. Alicandro G, Remuzzi G, Centanni S, Gerli A, La Vecchia C. Excess total mortality during the Covid-19 pandemic in Italy: updated estimates indicate persistent excess in recent months. Med Lav. 2022;113(2):e2022021. Published 2022 Apr 26. doi:10.23749/mdl.v113i2.13108

10. ISTAT. Aggiornamento della base dati di mortalità totale giornaliera comunale Gennaio-giugno e stima luglio 2022. Istituto Nazionale di Statistica. August 30, 2022. Available at: https://www.istat.it/it/archivio/274010 Accessed on: November 28, 2022.

11. Ferroni E, Giorgi Rossi P, Spila Alegiani S, et al. Survival of Hospitalized COVID-19 Patients in Northern Italy: A Population-Based Cohort Study by the ITA-COVID-19 Network. Clin Epidemiol. 2020;12:1337–1346. Published 2020 Dec 8. doi:10.2147/CLEP.S271763

12. Sousa PM, Trigo RM, Russo A, et al. Heat-related mortality amplified during the COVID-19 pandemic. Int J Biometeorol. 2022;66(3):457–468. doi:10.1007/s00484-021-02192-z

13. Scortichini M, De Sario M, de’Donato FK, Davoli M, Michelozzi P, Stafoggia M. Short-Term Effects of Heat on Mortality and Effect Modification by Air Pollution in 25 Italian Cities. Int J Environ Res Public Health. 2018;15(8):1771. Published 2018 Aug 17. doi:10.3390/ijerph15081771

14. Thom EC. The discomfort index. Weatherwise, 1959;12:59–60.

15. Uusküla A, Jürgenson T, Pisarev H, et al. Long-term mortality following SARS-CoV-2 infection: A national cohort study from Estonia. Lancet Reg Health Eur. 2022;18:100394. doi:10.1016/j.lanepe.2022.100394

16. Changes in the Effect of Heat on Mortality in the Last 20 Years in Nine European Cities. Results from the PHASE Project. Int J Environ Res Public Health. 2015;12(12):15567–15583. Published 2015 Dec 8. doi:10.3390/ijerph121215006

